# Long-read sequencing reveals novel isoform-specific eQTLs and regulatory mechanisms of isoform expression

**DOI:** 10.1101/2024.02.21.24302494

**Authors:** Yuya Nagura, Mihoko Shimada, Ryoji Kuribayashi, Hiroki Kiyose, Arisa Igarashi, Tadashi Kaname, Motoko Unoki, Akihiro Fujimoto

**Affiliations:** Department of Human Genetics, The University of Tokyo, Graduate School of Medicine, Tokyo, Japan; Department of Genome Medicine, National Centre for Child Health and Development, Tokyo, Japan

## Abstract

Genetic variations linked to changes in gene expression are known as expression quantitative loci (eQTLs). The identification of eQTLs provides a profound understanding of the mechanisms governing gene expression. However, prior studies have primarily utilized short-read sequencing techniques, and the analysis of eQTLs on isoforms has been relatively limited. In this study, we employed long-read sequencing technology (Oxford Nanopore) on B cells from 67 healthy Japanese individuals to explore genetic variations associated with isoform expression levels, referred to as isoform eQTLs (ieQTLs). Our analysis revealed 33,928 ieQTLs, with 69.0% remaining undetected by a gene-level analysis. Additionally, we identified ieQTLs that have significantly different effects on isoform expression levels within a gene. A functional feature analysis demonstrated a significant enrichment of ieQTLs at splice sites and specific histone marks, such as H3K36me3, H3K4me1, H3K4me3, and H3K79me3. Through an experimental validation using genome editing, we observed that a distant genomic region can modulate isoform-specific expression. Moreover, an ieQTL analysis and minigene splicing assays unveiled functionally crucial variants in splicing, which software-based predictions failed to anticipate. A comparison with GWAS data revealed a higher number of colocalizations between ieQTLs and GWAS findings compared to gene eQTLs. These findings highlight the substantial contribution of ieQTLs identified through long-read analysis in our understanding of the functional implications of genetic variations and the regulatory mechanisms governing isoforms.

## Introduction

Genetic variations associated with variations in gene expression are referred to as expression quantitative loci (eQTL). Identifying eQTL offers a deeper understanding of the mechanisms regulating gene expression [1,2,3]. Previous studies have unveiled that various genetic variations and genomic features, such as short insertions and deletions (indels), variations of microsatellites, transposable elements, chromatin states and histone marks, affect the pattern of gene expression [3,4,5,6]. Additionally, eQTL can help predict causative genes behind diseases and traits [1,7]. Genome-wide association studies (GWAS) have revealed huge amounts of genotype-phenotype associations [1,7]. However, a majority of GWAS hits were found in non-coding regions, posing a significant challenge in understanding their functional roles [1]. Moreover, closely linked variants within a region often exhibit a significant association due to linkage disequilibrium (LD), making it difficult to discern causative variants from multiple linked variants. Colocalizing GWAS variants with eQTL can aid in addressing these issues [1,7]. Previous studies have successfully identified causative variants and genes by comparing GWAS peaks with eQTLs [7–9]. Nevertheless, a colocalization analysis explains only a fraction of GWAS peaks [10], prompting the need for further eQTL analysis to comprehensively understand the biological mechanisms underlying diseases.

The analysis of isoforms has significant potential for expanding eQTL studies. Genes produce isoforms of various functions through alternative splicing, and isoforms are an important functional unit of genes. The impact of isoforms on diseases has been reported. For instance, splicing abnormalities are frequently observed in many Mendelian diseases, and changes in isoforms have been linked to the severity of infectious diseases [11,12]. Furthermore, functional isoforms have been identified in vascular smooth muscle cells [13], and oncogenic isoforms have been identified in hepatocellular carcinoma, breast cancer, and colon cancer [14–16]. These findings strongly indicate that isoforms play pivotal roles in the development and progression of diseases.

The recent advent of long-read sequencing technologies enables us to analyze full-length transcripts. The utilization of long-read sequencing for full-length transcript analysis enables a comprehensive observation of different isoforms within genes, significantly contributing to the understanding of gene expression [17,18]. While previous studies have utilized long reads to detect eQTLs, these efforts typically employed an allele-specific expression analysis on a limited number of samples or specific genes. Notably, a comprehensive eQTL analysis covering all isoforms across individuals within a population has yet to be conducted [17,18]. We propose that an expression analysis using long reads will unveil novel eQTLs, shed light on new regulatory mechanisms of gene expression, and facilitate the prediction of functional effects of GWAS variants.

In the present study, we sought genetic variations associated with isoform expression levels (referred to as isoform eQTLs, or ieQTLs). To achieve this, we performed an isoform analysis using long-read sequencing technology (Oxford Nanopore) on 67 immortalized B cell lines from healthy Japanese individuals [19]. Subsequently, we investigated the relationship between genetic variations and isoform expression levels. This analysis unveiled isoform-specific QTLs that have largely remained unreported in previous studies, underscoring the importance of a full-length transcriptome approach in human genetics.

## Material and Methods

### Samples and cDNA sequencing

Sixty-seven Japanese B cell samples from the 1000 Genomes Project were obtained from the Coriell Institute [19]. Institutional review boards (IRBs) at the University of Tokyo approved this work (2022121Ge). B cells were cultured in RPMI1640 medium (Nacalai) supplemented with 10% fetal bovine serum (FBS) and penicillin/streptomycin at 37°C in a 5% CO_2_ (Additional file 2: Table S1). Human embryonic kidney 293 (HEK293) cells were obtained from American Type Culture Collection and maintained in Dulbecco’s Modified Eagle Medium (DMEM) supplemented with 10% FBS and penicillin/streptomycin at 37°C in a 5% CO_2_. RNA extraction was carried out using the RNeasy Mini Kit (QIAGEN). Full-length cDNA was synthesized from 1 μg of total RNA using the SMARTer PCR cDNA Synthesis Kit (Takara) following the manufacturer’s instructions. Subsequently, primers for cDNA synthesis were digested with exonuclease I (NEB) at 37 °C for 30 minutes. After purification with Agencourt AMPure XP magnetic beads (Beckman Coulter), libraries were constructed using the Ligation Sequencing Kit (SQK-LSK109) (Oxford Nanopore) as per the manufacturer’s protocol. Sequencing was performed on a Flow Cell (R9.4) (Oxford Nanopore) using the MinION sequencer (Oxford Nanopore). Basecalling from FAST5 files was conducted using Guppy software (version 3.0.3) (Oxford Nanopore).

### Estimation of the isoform expression level

We estimated the expression levels of isoforms using our previously developed analysis pipeline, SPLICE [14]. Initially, SPLICE filtered out reads of low quality (average quality score < 15) and then aligned the remaining reads to the human reference genome (GRCh38) using minimap2 [20]. Subsequently, the mapped reads were annotated based on data from the GENCODE (version 28) and RefSeq (release 88) databases. The analysis provided the number of mapped reads corresponding to each isoform. These read counts were standardized to (number of reads)/(1M reads) for each isoform within each sample and were utilized as the expression level for each isoform. Subsequent analyses focused on genes located on autosomes.

### Polymorphism of the 67 Japanese individuals and annotation of variants

The genotype data for single nucleotide variants (SNVs) and short indels were sourced from the 1000 Genomes (1000G) database [19]. Single nucleotide polymorphisms (SNPs) and polymorphic indels were selected based on the following criteria using PLINK software [21]: call rate > 90%, p-value indicating departure from Hardy-Weinberg equilibrium > 0.001, and minor allele frequency (MAF) > 1%.

To assess the functional impact of SNPs and indels, we classified them into 9 categories: exon, intron, 5’ untranslated region (5’ UTR), 3’ UTR, 5’ splice sites (−2 bp∼+5 bp from the 5’ end of intron), splice donor sites, 3’ splice sites (−2 bp ∼ +5 bp from the 3’ end of intron), splice acceptor sites, and branch point regions [22].

The annotation of known regulatory regions was obtained from the Ensemble database, specifically from B cell line (GM12878) data [23]. The locations of super enhancers in GM12878 were retrieved from the dbSUPER database [24]. Positions in GRCh37 were converted to GRCh38 using the ‘liftover’ command in PLINK software. Information regarding histone modifications and transcription factor binding sites (TFBSs) in the B cell line was sourced from the Encyclopedia of DNA Elements (ENCODE) database. We filtered data from the ‘Experiment Matrix’ under the following conditions: Status - released, Perturbation - not perturbed, Organism - Homo sapiens, Genome assembly - GRCh38, and Available file types - bed narrowPeak. After filtering based on ‘Audit category’ (Red icon and orange icon), data from 280 files were utilized for this study [25].

### Identification of isoform eQTL (ieQTL)

We generated expression data for each isoform (Figure 1AB). Our primary goal in this study was to identify cis-eQTLs for each gene, focusing on genetic variations within the cis-window defined as 1 Mb from the transcription start site (TSS) and transcription end site (TES) of each gene. To analyze the association between isoform expression levels and genetic variations, we employed the matrix-eQTL package in R [26].

Next, we computed the number of tests within each cis-window to correct for multiple testing. Due to varying numbers of genetic variations and their high correlation resulting from LD, we determined the number of tests for each cis-window through LD pruning using PLINK software. LD pruning was conducted for variants with MAF > 3% using the following parameters: “window size in SNP” = 100, “The number of SNPs to shift the window at each step” = 5, and r^2^ > 0.5. Multiple test corrections for each cis-window was performed using the Bonferroni method based on the number of variants post LD pruning. Variants with Bonferroni-corrected p-values < 0.05 were deemed significant.

Due to the potential influence of LD, variants that are not causative may display low p-values. For genes or isoforms with more than 3 variants demonstrating Bonferroni-corrected p-values < 0.05, we employed the Finemap program [27] to identify potential causative variant candidates. Finemap computes the posterior probability that a set of variants is causal among all candidate variants. In this study, we considered sets with the highest posterior probability as causal variants, defining them as ieQTLs. Genes or isoforms with ≤ 2 variants showing Bonferroni-corrected p-values < 0.05 were not subjected to the Finemap analysis, and all variants within these cases were considered ieQTLs.

### Identification of gene eQTL

We also examined the expression levels of genes (Figure 1AB). This involved calculating the sum of read numbers for isoforms within each gene. The read counts for each gene were then standardized to (number of reads)/(1M reads) within each sample and utilized as the expression level for each gene. The process for identifying gene eQTLs was conducted similarly to the approach used for identifying ieQTLs.

### Comparison with previous studies

We conducted a comparison between our eQTL list and a previously conducted eQTL study, utilizing the eQTL data retrieved from the Genotype-Tissue Expression (GTEx) database [28]. Additionally, we also assessed the colocalization of our eQTLs with variants that showed significant associations in previous GWAS. We collected variants with associations from prior GWAS through the GWAS catalog database [29] and verified the presence of these SNPs in our eQTL list.

### Classification of ieQTL based on the effects on expression

Many genes express multiple isoforms, and each ieQTL can exert distinct effects on different isoforms within a gene (Figure 1AB). For instance, a SNP might positively regulate the expression of one isoform while negatively impacting another isoform (Figure 1C). To identify such ieQTL variations, we compared the β values derived from regression analyses of ieQTLs. Since the difference in β values conforms to a t-distribution, we employed a t-test to evaluate these differences. Two types of ieQTLs were categorized based on the analysis of β values. ieQTLs exhibiting significantly different effects (p-value < 0.01) on distinct isoforms within a single gene were termed ‘differential ieQTLs’ (Figure 1C). Among these, ieQTLs with β values displaying opposite signs were termed ‘opposite ieQTLs’ (Figure 1C).

**Figure 1.**
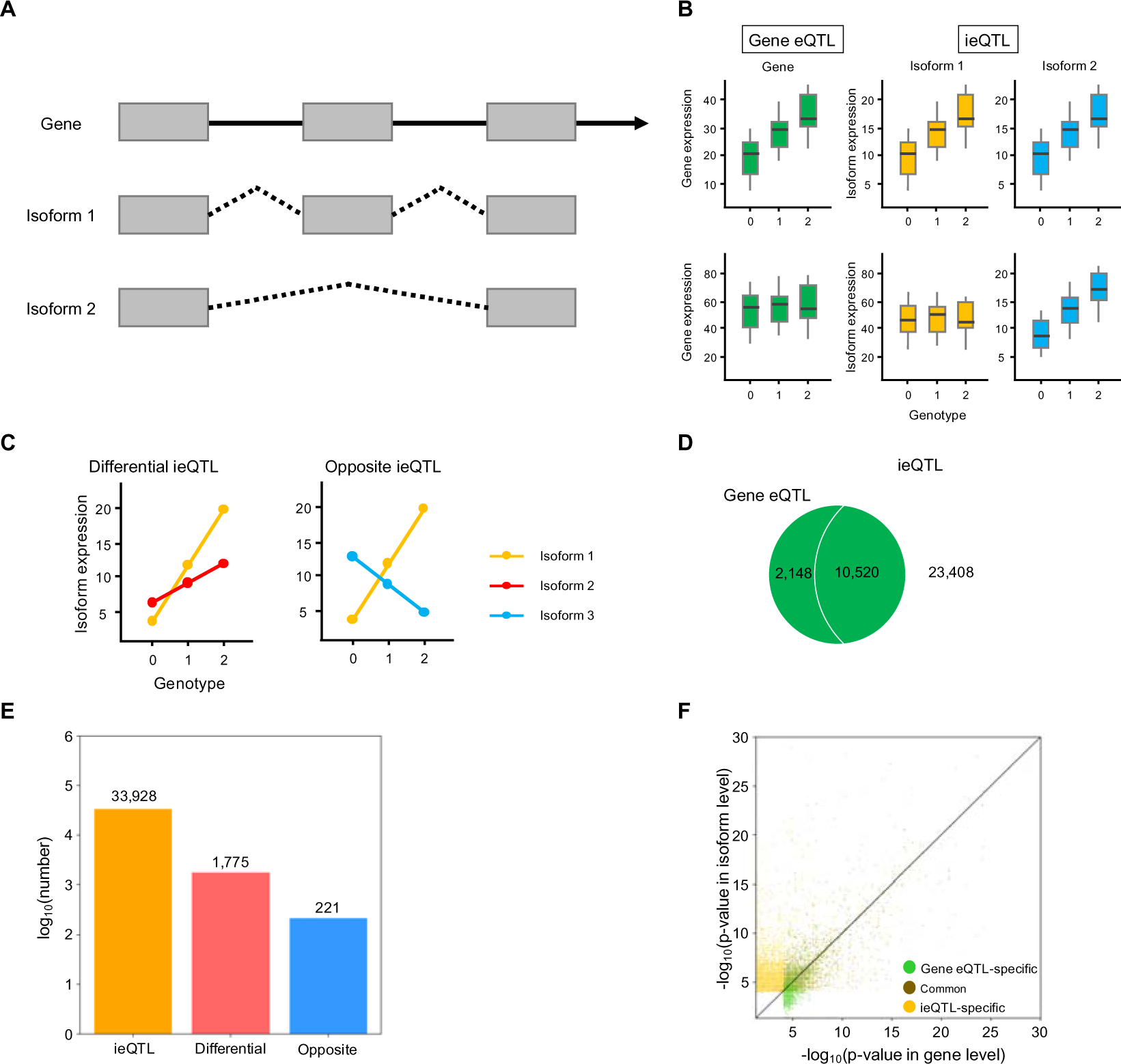
Isoform eQTLs (ieQTLs) and gene eQTLs. (A) Genes and isoforms. Multiple isoforms can be expressed from a single gene. Gray boxes, a solid line, and dotted lines indicate exons, introns, and splicing pattens, respectively. (B) Patterns of gene eQTLs (green) and ieQTLs (yellow and blue) by genotype (0, 1, and 2). Gene eQTLs are identified through the expression levels of entire genes, while ieQTLs are detected based on the expression levels of individual isoforms. (C) Differential ieQTLs and opposite ieQTLs. Differential ieQTLs are ieQTLs that exhibit significantly different effects on distinct isoforms. Opposite ieQTLs are differential ieQTLs with contrary effects on different isoforms. (D) Venn diagram of number of gene eQTLs (green) and ieQTLs (yellow) identified in this study. (E) Number of total ieQTLs (orange), differential ieQTLs (red), and opposite ieQTLs (blue) identified in this study. (F) Distribution of *p*-values (log_10_) from gene-level (green), common (brown), and isoform-level (yellow) analyses.

We conducted a multiple regression analysis on normalized β values of eQTLs using the lm function in R software. The independent variables considered were distance from TSS, conservation score (the average phyloP100way score within 200 bp), annotations (such as Activity-by-Contact (ABC) enhancer, CCTC-binding factor (CTCF) binding site, histone H3 trimethylation at lysine 36 (H3K36me3), SS3 (3’ splice site), SS5 (5’ splice site), TFBS, 3’ UTR, 5’ UTR, acceptor, branch, donor, enhancer, exon, intron, open chromatin region, promoter, promoter flanking region, super enhancer), and MAF. The step() function was utilized for the parameter selection.

The impact of genetic variations on splicing was estimated using the SpliceAI website [30].

### Enrichment analysis

To assess the potential enrichment of biological features among ieQTLs, we tallied counts for each regulatory feature observed in ieQTLs and compared them to counts found in non-causal variants. We categorized eQTLs into several groups and compared them as follows: (i) ieQTLs vs. no eQTLs, (ii) gene eQTLs vs. no eQTLs, (iii) common, which were common to both ieQTLs and gene eQTLs, vs. no eQTLs, (iv) differential ieQTLs vs. other ieQTLs, and (v) opposite eQTLs vs. other ieQTLs. We compared the regulatory features between these categorized groups using Fisher’s exact test. The analysis incorporated biological features outlined in ‘*Polymorphism of the 67 Japanese individuals and annotation of variants*’.

### Identification of motifs in isoforms affected by ieQTLs

We compared the major and minor isoforms influenced by ieQTLs. We defined the most abundant isoform as the major isoform and others as minor isoforms. The coding sequences of isoforms were translated into amino acids. Motifs were estimated for each isoform using HMMER software with default settings [31]. Amino acid sequences and predicted domains were then compared among the isoforms.

### Minigene splicing assay

To assess the impact of two SNPs (*interferon induced protein 44 like* (*IFI44L*) IVS2+3T/A (rs1333973) and *growth arrest specific 2* (*GAS2*) IVS1+3G/A (rs11026723)) on splicing, we conducted minigene splicing assays. For the analysis of rs1333973, a genomic segment 1753 bp in length encompassing the intron, 2nd exon, and 3rd exon of *IFI44L* gene was amplified and cloned into the pSPL3 plasmid vector (NovoPro Bioscience Inc.) (Additional file 1: Figure S1). The alternative allele was generated using the PrimeSTAR Mutagenesis Basal Kit (Takara Bio). Regarding the analysis of rs11026723, a genomic segment 589 bp in length and inclusive of the upstream flanking region, 1st exon, and intron of *GAS2* gene (NM_177553.2) was amplified and cloned into the pSPL3 vector. Subsequently, we deleted the 1st exon of the pSPL3 vector and the upstream flanking region of *GAS2* gene using the PrimeSTAR Mutagenesis Basal Kit. The alternative allele was generated using the same mutagenesis kit (Additional file 1: Figure S1).

Four vectors (pSPL3 with *IFI44L* IVS2+3T, *IFI44L* IVS2+3A, *GAS2* IVS1+3G, or *GAS2* IVS1+3A) were transfected into HEK293 cells utilizing FuGENE HD Transfection Reagent (Promega). Forty-eight hours post-transfection, RNA extraction from the cells was carried out using the RNeasy Mini Kit (QIAGEN). The total RNA underwent reverse transcription (RT) with the PrimeScript RT Reagent Kit with gDNA Eraser (Perfect Real Time) (Takara Bio). Subsequently, RT-PCR was performed (Additional file 2: Table S2), and the resulting amplicons were assessed via electrophoresis using a 2% agarose gel. Amplicons obtained through *IFI44L* IVS2+3T and *GAS2* IVS1+3G were purified using the QIAquick Gel Extraction Kit (QIAGEN) and subsequently sequenced using the Sanger sequencing method. A purified product derived from pSPL3 with *IFI44L* IVS2+3T contained multiple amplicons and underwent TA cloning with T-Vector pMD20 (Takara Bio). Eight colonies were selected for colony-PCR and subsequently sequenced using the Sanger sequencing method.

### Deletion with CRISPR-Cas9 system and examination of gene expression level

Our analysis identified numerous ieQTLs located outside of genes. To validate the impact of ieQTLs on gene expression, we induced a deletion in HEK293 cell lines using the Alt-R CRISPR-Cas9 System (Integrated DNA Technologies). We deleted the flanking region of rs11191660 (chr10:103343208 A/G), which was significantly associated with the expression level of an isoform (NM_032747.3) of the *ATP synthase membrane subunit K* (*ATP5MK*) gene. Two guide RNAs (gRNAs) were specifically designed for generating a deletion (Additional file 2: Table S3). Ribonucleoprotein complexes containing these gRNAs were formed and transfected into HEK293 cells using the Neon Transfection System (Thermo Fisher Scientific) with voltage = 1,150 V, width settings = 20, and pulse number = 2. The presence of the deletion in transfected cells was confirmed by PCR. The transfected cells were then separated into three wells and cultured. After 8-23 days post-transfection, DNA and RNA were extracted from the cells using the QIAamp DNA Mini Kit and RNeasy Mini Kit, respectively (QIAGEN).

The expression levels of the *ATP5MK* gene isoforms were assessed through RT-PCR quantification. Total RNA underwent reverse transcription using the PrimeScript RT Reagent Kit with gDNA Eraser (Perfect Real Time) (Takara). PCR primers were designed to amplify each isoform (Additional file 2: Table S2). The resulting cDNA was employed for quantitative PCR (qPCR) with KAPA SYBR Fast qPCR KIT (KAPA Biosystems) using CFX-Connect Real-Time System (Bio-Rad), utilizing *Actin beta* (*ACTB*) as an internal control (Additional file 2: Table S2). The biological triplicate samples with and without deletions were used for real-time PCR in technical triplicate, and the relative quantification (RQ) values were calculated. Statistical significance of the RQ values between cells with and without the deletion was determined using the Student’s t-test.

## Results

### cDNA sequencing and estimation of expression level

We sequenced cDNA obtained from 67 Japanese B cell lines. The average data yield was 15.8 Gbp, and the average number of reads was 13.1 million (Additional file 2: Table S1, S4). The average read length was 1222.6 bp. Subsequently, reads were aligned to the human reference genome (GRCh38) using minimap2 software [20]. Annotation of the reads and detection of the isoform expression levels were performed using SPLICE software [14]. Among the genes in autosomes, a total of 43,581 isoforms (representing 15,353 genes) were expressed (Additional file 2: Table S4). Of these, 6,832 were not identified in the Refseq or GENCODE databases; hence, they were classified as novel.

### Identification of ieQTL and gene eQTL

Based on the expression levels of each isoform and genotype data, we detected ieQTLs. Additionally, we computed the total number of reads mapped to each gene to identify gene eQTLs. Our analysis revealed 33,928 ieQTLs affecting 12,202 isoforms (from 7,683 genes) and 12,668 gene eQTLs (from 4,344 genes) (Figure 1D, Additional file 2: Table S5). Out of these, 10,520 were common to both ieQTLs and gene eQTLs, 23,408 were ieQTL-specific, and 2,148 were specific to gene eQTLs (Figure 1D). ieQTLs accounted for 0.45% of all analyzed variants (7,484,843 non-redundant variants within 1 Mbp of cis-windows). Among the ieQTLs, 1,775 were identified as differential ieQTLs, and 221 exhibited opposite effects (Figure 1E, Additional file 2: Table S6, 7). A subsequent comparison of p-values between ieQTLs and gene eQTLs for each variant revealed that numerous variants displayed lower ieQTL p-values than gene eQTL p-values, suggesting that ieQTLs are distinct and not statistical artifacts arising from gene eQTL identification (Figure 1F). A comparison of our eQTL list with GTEx eQTL and sQTL studies using lymphocyte samples demonstrated that 86.7% and 88.2% of the identified eQTLs were not detected, respectively (Additional file 1: Figure S2).

Our analysis identified a larger number of ieQTLs compared to gene eQTLs. In this analysis, we conducted multiple test corrections based on the number of variants within 1 Mbp from genes. As the total number of isoforms exceeded the number of genes, the number of tests became greater for ieQTLs (n = 43,581) than for gene eQTLs (n = 15,353), potentially contributing to the higher count of ieQTLs. To examine this hypothesis, we adjusted the p-values of the ieQTL analysis using the Bonferroni correction considering the number of isoforms and variants for each gene. Since the expression levels of isoforms within the same gene may not be entirely independent, this correction might be overly conservative. Despite this correction, we still identified 25,205 ieQTLs, of which 15,422 were not identified as gene eQTLs. These findings indicate that the ieQTL analysis revealed a larger number of regulatory variants compared to the gene-based analysis.

### Features of ieQTLs and gene eQTLs

To explore functional features associated with ieQTLs, we conducted an enrichment analysis for 27 functional categories (gene location, regulatory regions, histone modifications, and TFBSs) across three distinct groups: ieQTLs exclusively found in the ieQTL group (ieQTL-specific), those exclusively present in the gene eQTL group (gene eQTL-specific), and those commonly found in both the ieQTL and gene eQTL groups (common) (Figure 2A-C, Additional file 2: Table S8). Following multiple test corrections, 26 categories exhibited significance within the ieQTL-specific group. As expected, the ieQTL-specific group showed a significant enrichment in acceptor sites, donor sites, 3’ splice sites, and 5’ splice sites with high odds ratios (OR) (acceptor site: p-value=1.3×10^−8^, OR=6.4; donor site: p-value=3.1×10^−6^, OR=5.1; 3’ splice site: p-value=7.8×10^−14^, OR=5.4; 5’ splice site: p-value=3.8×10^−23^, OR=4.7). Additionally, significant enrichments were observed in promoters, branchpoints within introns, and super enhancers (promoter: p-value=2.2×10^−198^, OR=2.5; branchpoint: p-value=1.7×10^−13^, OR=2.2; super enhancer: p-value=4.6×10^−23^, OR=2.1). Within the gene eQTL-specific group, 14 categories showed significant enrichment. Promoters exhibited the highest odds ratio, followed by 3’ UTRs, TFBSs, exons, and 5’ UTRs (promoter: p-value=9.4×10^−40,^ OR=3.4; 3’ UTR: p-value=2.9×10^−11^, OR=2.0; TFBS: p-value=1.9×10^−18^, OR=1.8; exon: p-value=1.9×10^−11^, OR=1.8; 5’ UTR: p-value=9.9×10^−5^, OR=1.6). Subsequently, we compared the number of variants within each category between the ieQTL-specific group and the gene eQTL-specific group (Figure 2A-C, Additional file 2: Table S9). The proportion of eQTLs present in super enhancer and H3K36me3 regions was significantly higher within the ieQTL-specific group (super enhancer: p-value=5.1×10^−5^, OR=5.1; H3K36me3: p-value=1.6×10^−5^, OR=1.3), while the proportion in promoters was greater in the gene eQTL-specific group (p-value=6.8×10^−4^, OR=0.74).

**Figure 2.**
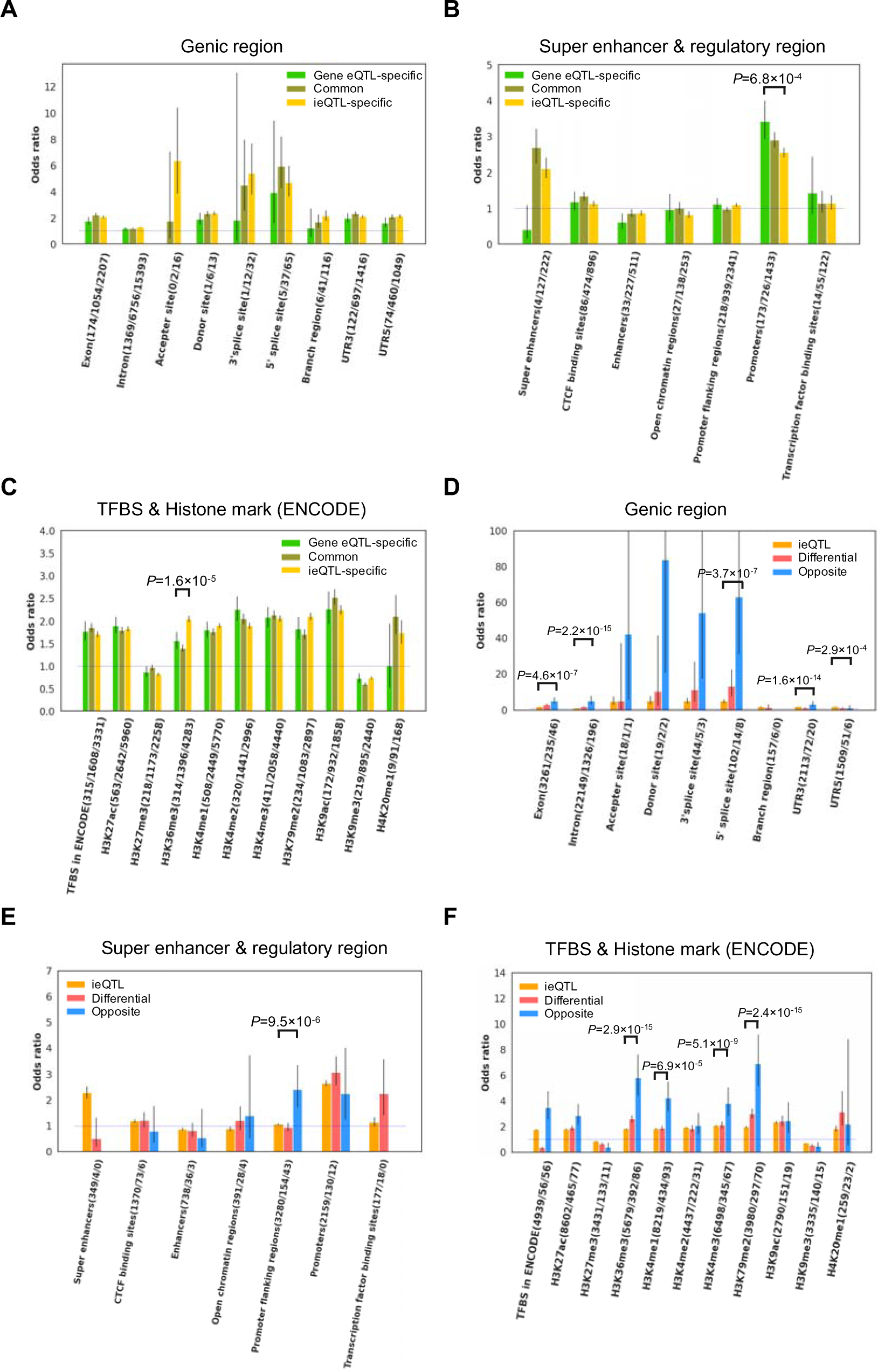
Functional enrichment analysis of eQTLs eQTLs were categorized into gene eQTL-specific (green), common (brown), and ieQTL-specific (yellow) groups. Functional enrichment was assessed for gene region annotations (A), super enhancers and regulatory regions (B), as well as transcription binding sites (TFBS) and major active and suppressive histone marks (ENCODE), which could associate with splicing (C). eQTLs were categorized into gene eQTL-specific (orange), differential ieQTL (red), and opposite ieQTL (blue) groups. A functional enrichment analysis was conducted for gene region annotations (D), super enhancers and regulatory regions (E), and TCFB and histone marks (F). The Y-axis indicates the Odds ratio. Presented are odds ratios and 95% confidence intervals.

We conducted an enrichment analysis for 126 TFBSs (Additional file 2: Table S10). Following multiple test corrections, 115 TFBSs exhibited a significant enrichment in the ieQTL-specific group, while 78 were significantly enriched in the gene eQTL-specific group. The proportion of CAMP responsive element binding protein 1 (CREB1) binding sites showed a significant difference between the ieQTL-specific group and the gene eQTL-specific group (Additional file 2: Table S11). These findings suggest that the regulatory mechanism governing isoform expression differs, at least partially, from that of genes (Figure 2A-C).

### Features of differential eQTLs and opposite eQTLs

We then proceeded with an enrichment analysis within the differential ieQTL group and the opposite ieQTL group (Figure 2D-F, Additional file 2: Table S12). After multiple test corrections, 16 categories, including 3’ and 5’ splice sites, exhibited significances in the differential ieQTL group (3’ splice site: p-value=1.1×10^−4^, OR=11.2; and 5’ splice site: p-value=8.9×10^−12^, OR=13.4). In the opposite ieQTL group, 16 categories were significant. Among these, 3’ and 5’ splice sites in the differential ieQTL group displayed particularly high odds ratios (donor sites: p-value=2.8×10^−4^, OR=84.0; 3’ splice site: p-value=2.8×10^−5^, OR=54.4; and 5’ splice site: p-value=1.7×10^−12^, OR=63.1). We subsequently compared the variants within each category between the differential group and other ieQTLs, as well as between the opposite ieQTL group and other ieQTLs (Figure 2D-F, Additional file 2: Table S13). The proportion of variants in exons, introns, and 5’ splice sites was significantly higher in the differentiated ieQTL group than in other ieQTLs (Figure 2D). Moreover, several histone marks, including H3K36me3, H3K4me1, H3K4me3, and H3K79me3, were significantly enriched in opposite ieQTLs compared to other ieQTLs (Figure 2F). Additionally, we conducted an enrichment analysis for 126 TFBSs (Additional file 2: Table S14, S15). Of these, 69 and 5 TFBSs were significantly enriched in the differential and opposite groups, respectively.

### Effects of ieQTLs

We analyzed the distribution of eQTLs (ieQTL-specific, common, and gene eQTL-specific groups) relative to the gene body (Figure 3A). The gene eQTL-specific group displayed a singular peak distribution, centered around the TSS. Conversely, the distribution of the ieQTL-specific group exhibited an additional peak around the TES. The common group showed an intermediate pattern between the gene eQTL-specific group and ieQTL-specific group. These observed patterns are consistent with findings from a prior study [17].

**Figure 3.**
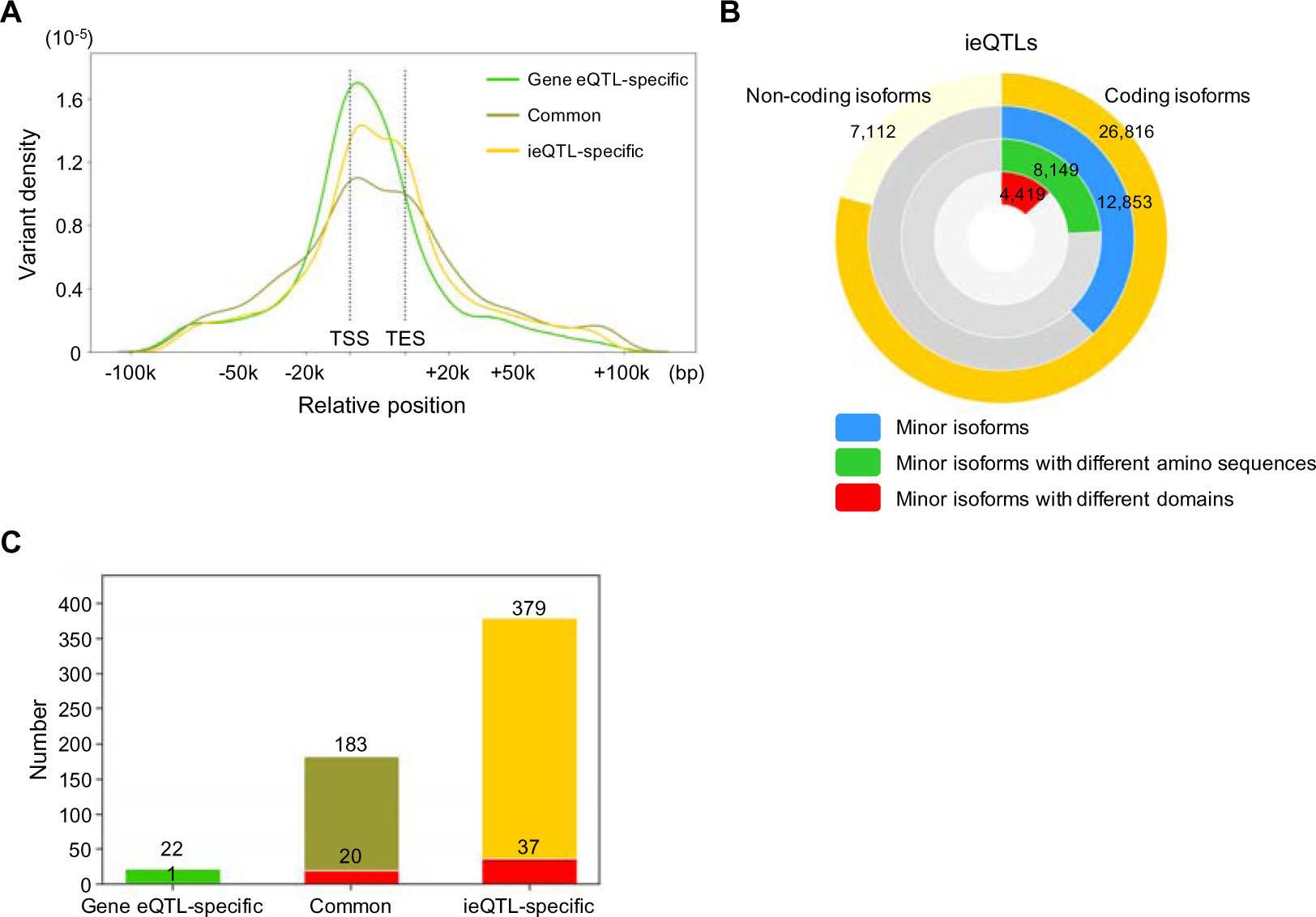
Effects of ieQTLs. (A) Distribution of gene eQTL-specific (green), common (gray), and ieQTL-specific (yellow) variants relative to the gene body. Gene size was adjusted to 25,000 bp, which is the average gene size. TSS, transcription start site; TES, transcription end site. (B) Comparison of amino acid sequences and functional motifs between major isoforms and minor isoforms affected by ieQTLs. (C) Number of GWAS hits found in gene eQTL-specific (green, n = 22), common (brown, n = 183), and ieQTL-specific (yellow, n = 379). Red bars indicate B-cell or autoimmune diseases in GWAS hits.

To examine the factors influencing the effect of eQTLs, we conducted a multiple regression on the effect sizes (β values) while considering various annotations of eQTLs (regulatory features, distance from TSS, MAF, and average conservation score within 200 bp) (Additional file 2: Table S16). We specifically examined genes with five or more isoforms. Following the selection of independent variables, eight features (conservation, ABC enhancer, 5’ splice site, donor site, enhancer, exon, intron, and MAF) were associated with the effect sizes of the ieQTL-specific group. Conversely, two factors (CTCF binding site and MAF) were associated with the effect sizes within the gene eQTL-specific group (Additional file 2: Table S16).

We subsequently examined differences in amino acid sequences and motifs in isoforms affected by ieQTLs (Figure 3B, Additional file 2: Table S17). Specifically, we focused on ieQTLs that influenced coding genes. Within this analysis, we categorized isoforms based on their expression levels, designating the most abundant isoform as the major isoform and others as minor isoforms. We specifically selected minor isoforms influenced by ieQTLs and compared their amino acid sequences with those of the major isoforms (Figure 3B). Among the 33,928 ieQTLs identified, 26,816 affected minor isoforms. Among these, 30.4% (8,149/26,816) displayed a distinct amino acid sequence from the major isoforms, while 16.5% (4,419/26,816) exhibited different functional domains. These findings suggest that ieQTLs have the potential to induce functional alterations.

Combining eQTL and GWAS data has proven effective at predicting causative variants [7]. Hence, we examined whether the identified QTLs corresponded to disease-associated variants previously reported in GWAS studies. We found that 1.0% of the gene eQTL-specific group (22/2,148), 1.6% of the ieQTL-specific group (379/23,408), and 1.7% of the common eQTLs (183/10,520) were present among GWAS-associated variants (Figure 3C, Additional file 2: Table S5). Interestingly, the proportion of GWAS variants was higher in the ieQTL-specific group than in the gene eQTL-specific group (Fisher’s exact test, p-value = 0.036, OR=1.6). This finding suggests that an ieQTL analysis aids in identifying novel mechanisms underlying diseases.

### Prediction of impact on splicing and validation by minigene splicing assay

Within the ieQTL-specific group, there were 146 variants in 3’ or 5’ splice sites, and 37 variants in donor or acceptor sites (Additional file 2: Table S5). Among these ieQTLs, we selected 94 variants present in affected genes and predicted their functional impact on splicing using SpliceAI software (Additional file 2: Table S18) [30]. Out of these, 18 eQTLs had a score ≥ 0.8 (“high precision” score by SpliceAI), and 12 had a score between 0.5 and 0.8 (“recommended” score by SpliceAI).

To investigate whether variants with low SpliceAI scores affect splicing, we chose two variants in the 3rd position of introns with low scores (*IFI44L* IVS2+3T/A and *GAS2* IVS1+3G/A) for minigene splicing assays (Figure 4A-I, Additional file 2: Table S18). *IFI44L* IVS2+3A/T (rs1333973) is in the 3rd position of intron2 of the *IF144L* gene. In the eQTL analysis, *IFI44L* IVS2+3T/A (rs1333973) did not impact the overall expression level of *IFI44L* gene but exhibited opposing effects on isoforms (Figure 4D). NM0068020.3 and XM_0115405392 showed the highest expression in A/A individuals, while XM_005270391.3 and XM_017000120.1 showed the highest expression in T/T individuals. The minigene splicing assay showed that *IFI44L* IVS2+3T/A affects the splicing pattern. *IFI44L* IVS2+3T (Figure 4E lane 2) reduced the amount of longer fragments (exon2-3) and generated short fragments caused by the skipping of exon 2 and splicing aberration.

**Figure 4.**
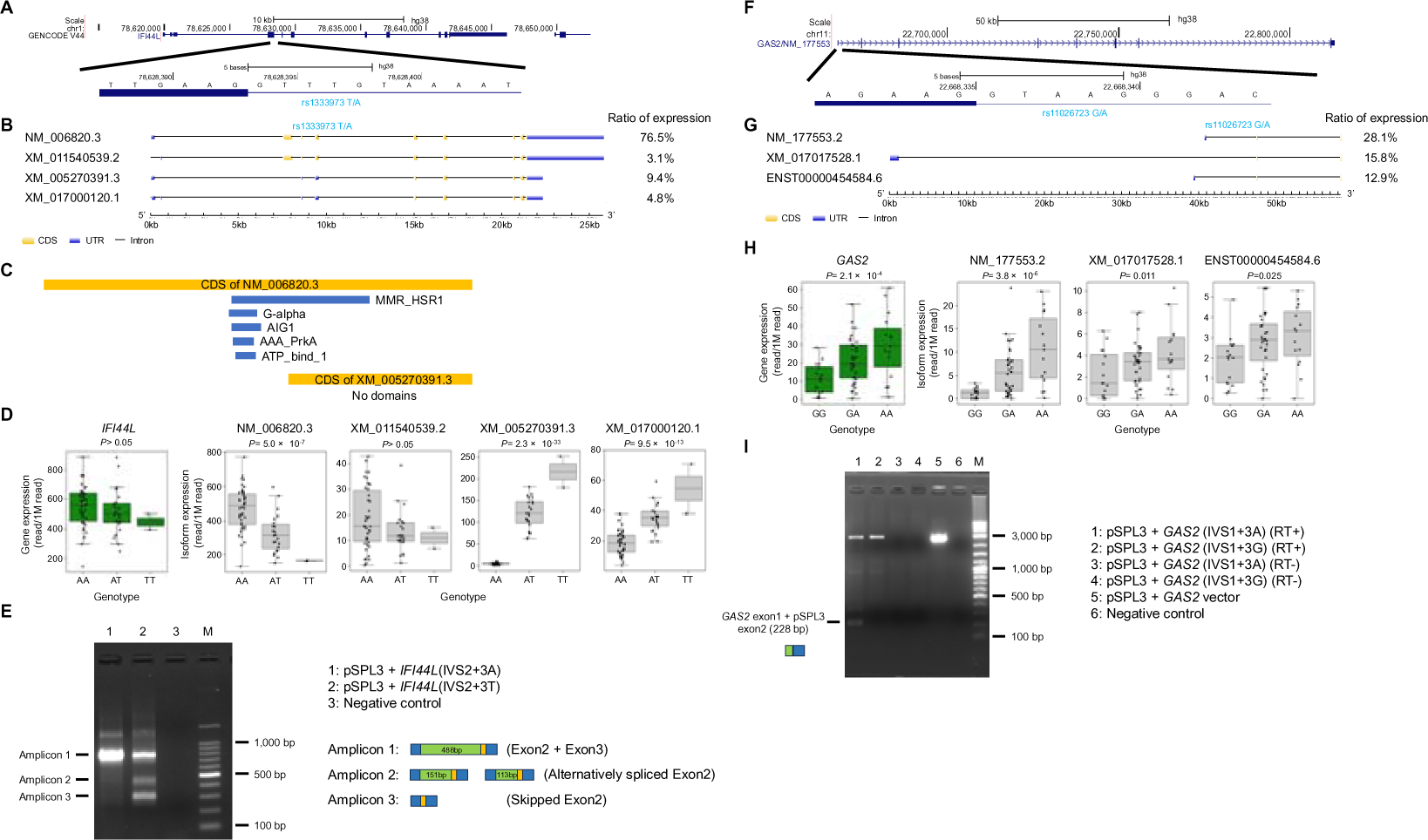
Minigene splicing assay for *IFI44L* and *GAS2* (A) Structure of *IFI44L* gene and the surrounding area of rs1333973. Exons and introns are depicted with bold and thin lines, respectively. This figure was obtained from the UCSC genome browser. (B) Isoforms of *IFI44L*. Isoform structures are depicted using GSDS 2.0. (C) Motif prediction for NM_006820.3 and XM_005270391.3 using HMMER software. MMR_HSR1, G-alpha, AIG1, AAA-PrkA, and ATP_bind_1 domains/motifs were predicted in CDS of NM_006820, while no domain/motif was predicted in CDS of XM_005270391.3. (D) Boxplot displaying gene expressions in the 67 Japanese B cell lines. The expression levels of the entire *IFI44L* gene (left) and four isoforms are presented by genotypes (AA (n = 42), AT (n = 23), and TT (n = 2)). (E) Agarose gel electrophoresis of RT-PCR products expressed from *IFI44L* (exon 2 and exon 3) minigenes in HEK293 cells. M: 100 bp ladder marker. Lane 1: pSPL3 + *IFI44L*(IVS2+3A). Lane 2: pSPL3 + *IFI44L*(IVS2+3T). Lane 3: Negative control. Blue box: exon of pSPL3 plasmid, green box: exon 2 of *IFI44L*, orange box: exon 3 of *IFI44L*. (F) Structure of *GAS2* gene and the surrounding area of rs11026723. Exons and introns are shown using bold and thin lines. This figure was obtained from the UCSC genome browser. (G) Isoforms of *GAS2*. Isoform structures are depicted using GSDS 2.0. (H) Boxplot showing gene expressions in the 67 Japanese B cell lines. The expression levels of the entire *GAS2* gene (left) and isoforms are presented by genotypes (GG (n = 16), GA (n = 36), and AA(n = 15).). (I) Agarose gel electrophoresis of RT-PCR products expressed from *GAS2* (exon 1) minigenes in HEK293 cells. In this experiment, a modified pSPL3 plasmid was utilized (Additional file 1; Figure S1). M: marker (DNA ladder One (Nacalai)). Lane 1: pSPL3 + *GAS2* (IVS1+3A) (RT+). Lane 2: pSPL3 + *GAS2* (IVS1+3G) (RT+). Lane 3: pSPL3 + *GAS2* (IVS1+3A) (RT-). Lane 4: pSPL3 + *GAS2* (IVS1+3G) (RT-). Lane 5: pSPL3-GAS2 plasmid. A purified plasmid was used as the PCR template. Lane 6: Negative control. RT: Reverse transcription. The size of the upper band in lanes 1 and 2 was the same as the amplicon in lane 5, suggesting the presence of an unspliced transcript. Blue box: exon of pSPL3 plasmid, green box: exon 1 of *GAS2*.

*GAS2* IVS1+3G/A is located in the 3rd position of the first intron of *GAS2* gene and primarily associated with the expression level of one isoform (NM_177553.2) (Figure 4H). In the minigene splicing assay, *GAS2* IVS1+3A generated the *GAS2* short fragment (228 bp) through splicing (Figure 4I lane 1), whereas *GAS2* IVS1+3G only produced the unspliced fragment (Figure 4I lane 2). This result suggests that *GAS2* IVS1+3G/A strongly affects the splicing.

### Experimental validation of influence of a SNP on isoform-specific expression

Our analysis revealed numerous ieQTLs located at a distance from the target gene, implying the involvement of regions outside the gene in regulating isoform expression (Figure 3A). To directly establish their functional impact, we induced a deletion in HEK293 cells (Additional file 1: Figure S3) and assessed its effect on gene expression levels using quantitative real-time RT-PCR. For this experiment, we specifically targeted regions containing a variant rs11191660 (Figure 5A). While rs11191660 showed a significant association with the expression level of an isoform (NM_032747.3) of the *ATP5MK* gene (A/A individuals showed the highest expression), its association with other isoforms was not statistically significant (ENST00000369825.5) or opposite (NM_001206426.1 and NM_001206427.1) (Figure 5BC). We generated a deletion of 686 bp (chr10:103,342,876-103,343,562) within the region harboring rs11191660 using the CRISPR-Cas9 system in HEK293 cells (Additional file 1: Figure S3, Additional file 2: Table S2). Subsequently, qPCR was performed (Additional file 2: Table S2). The results demonstrated that cells with the deletion exhibited significantly lower gene expression levels for NM_032747.3 (t-test p-value = 1.5×10^−6^) (Figure 5D). In contrast, the influence on the expression levels of other isoforms (NM_001206426.1 and NM_001206427.1) was either weaker or insignificant (Figure 5D). These findings strongly suggest that the region containing rs11191660 plays a functional role in regulating isoform expression.

**Figure 5.**
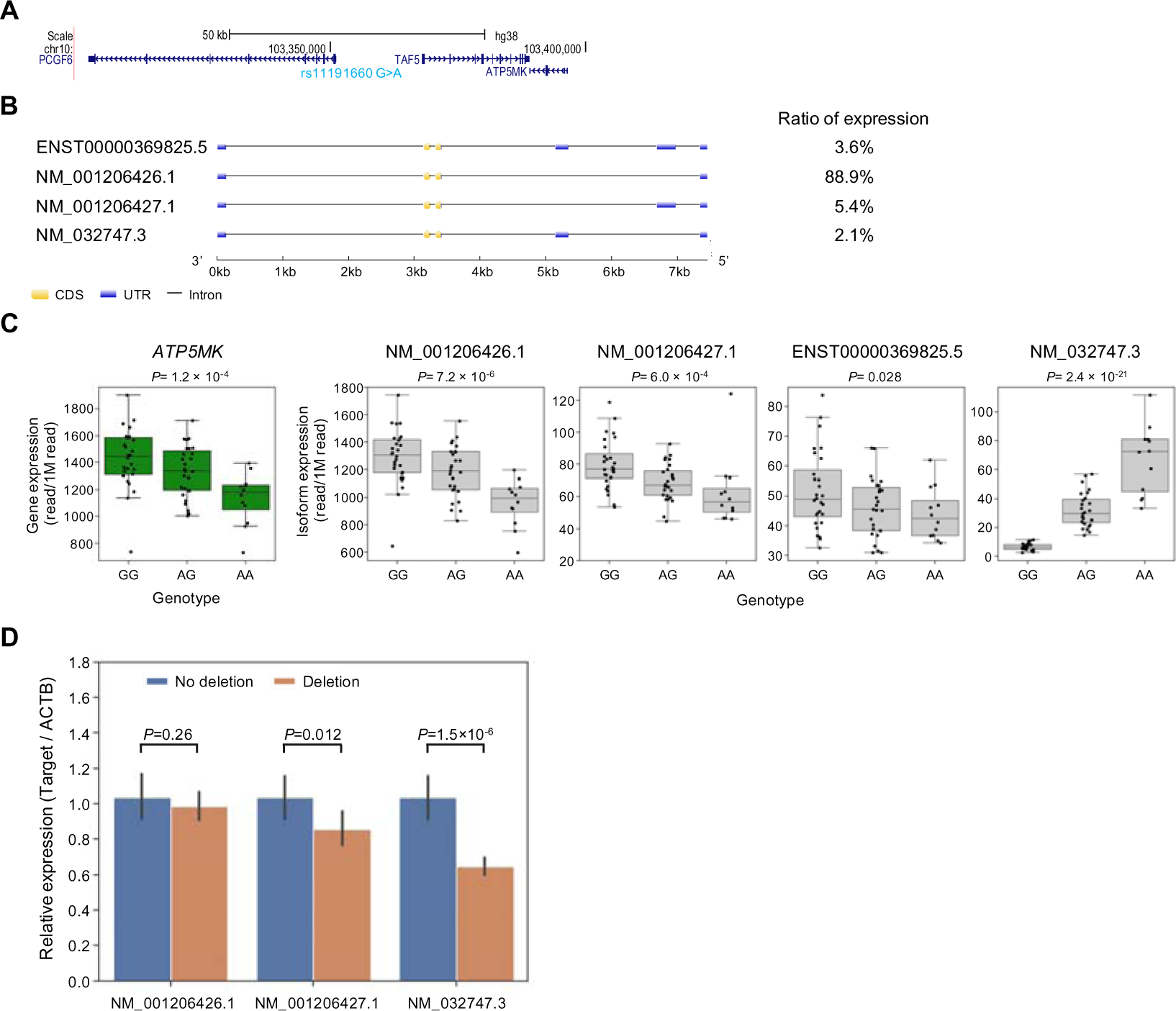
Effects of a deletion in HEK293 on gene expression. (A) Location of rs11191660 within the *ATP5MK* gene. (B) Four isoforms, ENST00000369825.5, NM_001206426.1, NM_001206427.1, NM_032747.3, of *ATP5MK*. Isoform structures are depicted using GSDS 2.0. (C) Boxplot displaying gene expressions in the 67 Japanese B cell lines. The expression levels of the entire *ATP5MK* gene (left) and isoforms are presented by genotype (GG (n = 28), GA (n = 27), and AA (n = 12)). (D) Comparison of gene expression levels between HEK293 cells with and without the chr10:103,342,876-103,343,562 deletion. *ACTB* was used an internal control. The y-axis shows relative expression of the target variants in cells with the deletion compared to cells with no deletion, which is calculated as 1.0. A significant difference was observed in the gene expression levels of NM_032747.3 between cells with and without the deletion. Bar graphs are shown as the mean value ± standard error (s.e.). P-values were obtained by the student’s t-test.

## Discussion

The expression patterns of isoforms vary among tissues and between cancerous and non-cancerous tissues [14,15,16,17,18]. These variations likely lead to functional changes in genes and regulate diverse biological processes [14,15,16,17,32]. Despite the intriguing nature of the regulatory mechanisms governing isoform expression, this area remains understudied. To uncover ieQTLs, we conducted an eQTL analysis using long-read sequencing technology, revealing 33,928 ieQTLs, of which 69.0% (23,408/33,928) were undetected by a gene-level analysis (Figure 1D). Upon examining the expression changes, we identified 1,775 differential ieQTLs that were difficult to detect via the gene-level analysis (Figure 1E). Additionally, we identified 221 opposite ieQTLs, representing an extremely distinctive pattern. These findings strongly indicate that an ieQTL analysis unveils a larger number of eQTLs compared to a gene-based eQTL analysis.

To explore the regulatory mechanisms of isoforms, we conducted an enrichment analysis (Figure 2). While the majority of the results were similar between gene eQTLs and ieQTLs, promoters were significantly overrepresented in the gene eQTL-specific group, whereas acceptor sites, 5’ splice sites, 3’ splice sites, and H3K36me3 were notably overrepresented in the ieQTL-specific group (Figure 2A-C). This finding suggests that promoters play a more substantial role in overall gene expression, while variants in splicing motifs and H3K36me3, which is a marker generally accumulated on the gene-body of active genes, influence isoform expression. The enrichment analysis of opposite ieQTLs showed very strong enrichment in 5’ and 3’ splice sites (Figure 2D), suggesting that the most opposite QTLs can be caused by variants in splicing sites. Additionally, several histone marks, including H3K36me3, H3K4me1, H3K4me3, and H3K79me2, showed a significant enrichment in opposite ieQTLs (Figure 2F). These histone marks are known as splicing-associated chromatin signatures [33]. Although a previous large-scale eQTL study using short reads showed a higher enrichment of H3K36me3 in splicing QTL (sQTL) than in eQTL, other histone marks were not reported [2]. Thus, our long-read eQTL study will contribute to a deeper understanding of the mechanisms regulating isoform expression.

In addition to variants within genes, our analysis showed that 82.5% of ieQTLs (28,004/33,928) were outside genes (Figure 3A, Additional file 2: Table S5). A multiple regression analysis revealed that variations in ABC enhancers and enhancers could account for differences in the effect size of ieQTLs (Additional file 2: Table S16). Furthermore, our genome-editing experiment indicated that a region approximately 60 kbp distant from *ATP5MK* gene can influence isoform expression (Figure 5). Although the functional mechanism behind this expression regulation remains unclear, the HaploReg website predicts that rs11191660 G/A can alter the activator protein 1 (AP-1) binding motif (Additional file 1: Figure S4) [34]. AP-1 has been associated with long-range enhancer interactions, which control transcription [35]. Consequently, this SNP might influence isoform expression through modulation of the 3D chromatin structure. This result suggests that intergenic regions, including enhancers, can regulate isoform expression.

Our analysis revealed isoform-specific expression changes caused by variants in splice sites (Figure 4). We selected two variants at the 3rd position of introns for experimental validation (*IFI44L* IVS2+3T/A and *GAS2* IVS1+3G/A). Although a prior study suggested that *IFI44L* IVS2+3T/A induces splicing alterations [36], experimental validation has not been conducted. Furthermore, SpliceAI software predicted a low functional impact for the SNP (Δscores = 0.34) (Additional file 2: Table S18). Therefore, conducting functional experiments to determine the causality of this SNP is needed. In the minigene assay, *IFI44L* IVS2+3T/A caused a change in the splicing pattern, resulting in the generation of shorter isoforms, consistent with the data analysis (Figure 4D). Another variant (*GAS2* IVS1+3G/A), which also exhibited a low SpliceAI Δscore (Δscores = 0.35), significantly influenced the splicing pattern (Figure 4H, Additional file 2: Table S18). In the minigene assay, splicing did not occur in the transcript from *GAS2* IVS1+3G vector, indicating a crucial role for the 3rd position in splicing. Considering its high evolutionary conservation (Additional file 1: Figure S4), this site likely holds significant functional importance. Because this variant affected the expression level of only one isoform (Figure 4H), this pattern should be difficult to detect using short-read sequencing. Overall, we believe that the functional impact of variants in splice sites cannot be accurately predicted solely by software and short-read analysis. This underscores the significant advantage of eQTL analysis employing long-read sequencing.

Through our analysis, we identified genes affected by eQTLs. Among these, *ACTB*, which has been commonly utilized as an internal control in expression analyses, exhibited the highest β values (Additional file 2: Table S5). Although the normalized β value (normalized contribution of the eQTL to the total expression of *ACTB*) for this ieQTL is not high, it could potentially contribute to differences in the expression levels of *ACTB* among individuals. This finding suggests that caution may be necessary when using *ACTB* as a control in certain contexts (Additional file 2: Table S5).

Integrating eQTL and GWAS contributes significantly to understanding the biological mechanisms behind diseases and traits. Our investigation revealed 584 eQTLs present in GWAS results (Figure 3C). Among these, 64.9% (n=379) were exclusively identified within ieQTLs, which is difficult to detect by a gene-level analysis. This result strongly suggests that a colocalization analysis of ieQTLs could uncover novel causative genes in GWAS. A recent study reported that only a small fraction of GWAS peaks coincide with eQTLs, which has raised concerns about a ‘missing regulation’ issue [37]. The consideration of ieQTLs helps to identify a larger number of regulatory variants and could contribute to addressing a part of this concern.

Moreover, the ieQTL analysis suggested a contribution of isoforms to disease. For example, an ieQTL against an isoform (XM_005270391.3) of *IFI44L* gene, which was affected by ieQTL is in 5’ splice site (Figure 4), was reported to be associated with immune responses to measles vaccine [38]. This splicing affects the expression level of two isoforms, of which one isoform (XM_005270391.3) lacks functional domains (Figure 4C) and whose changes should strongly affect the function of this gene. *ATP5MK* has associations with various phenotypes, such as height, smoking status, and risk of systemic lupus erythematosus, indicating that this isoform-specific regulation may have clinical importance [39–41] (Figure 5). An ieQTL (chr12:112,919,404) against an isoform (NM_016816.3) of the *2’-5’-oligoadenylate synthetase 1* (*OAS1*) gene, which is related to innate immunity, was reported to be associated with SLE (Systemic lupus erythematosus). NM_016816.3 encode different amino acid sequences from major isoforms (ENST00000452357), and the differences may have a functional effect (Additional file 2: Table S17). Moreover, our ieQTL analysis identified different genes from GWAS-predicted causative genes. For example, one variant (chr14:35,292,469) was reported to associate with allergy diseases (asthma, hay fever and eczema) in the GWAS database, and *proteasome 20S subunit alpha 6* (*PSMA6*) was a predicted to be a causative gene. However, that variant was an ieQTL (XM_005267782.3) in *protein phosphatase 2 regulatory subunit B gamma* (*PPP2R3C*) gene. Because *PPP2R3C* encodes a subunit of protein phosphatase 2 (PP2A), which is related to inflammatory responses (PP2A) [42], it may be another plausible causative gene of this association.

In this study, we examined ieQTLs using long-read sequencing technology. Consequently, our eQTL analysis uncovered numerous ieQTLs, pinpointing isoform-specific expression changes related to splice sites, histone marks, and enhancers. While the exact role of enhancers requires further clarification, our functional analysis revealed that regions distant from genes can regulate isoform expression. By combining our eQTLs with GWAS variants, we propose novel candidates for disease causation and mechanisms. We believe that delving into ieQTLs through long-read analyses will contribute significantly to our comprehension of the functional implications of genetic variations and the regulatory mechanisms governing isoforms.

## URLs

ENCODE database https://www.encodeproject.org/matrix/?type=Experiment&control_type!=*&status=released&perturbed=false

eQTL from GTEx database https://www.gtexportal.org/home/

GWAS catalog database https://www.ebi.ac.uk/gwas/

HaploReg website https://pubs.broadinstitute.org/mammals/haploreg/haploreg.php

SpliceAI https://spliceailookup.broadinstitute.org

GSDS (Gene Structure Display Server) 2.0 http://gsds.gao-lab.org/index.php

Coriell Institute https://www.coriell.org

## Data Availability

All data produced in the present work are contained in the manuscript

## Declarations

### Availability of data and materials

Nanopore MinION long-reads sequencing data is available from the Japanese Genotype-phenotype Archive (JGA) database. Accession number: XXXXXXXXX.

### Authors’ contributions

AF designed the study. YN, MS, RK and AF performed the computational analyses. YN, MS, AI and AF performed the experiments. YN, TK, UM and AF interpreted the results. YN and AF wrote the manuscript. All authors approved the final manuscript.

## Acknowledgements

The super-computing resource was provided by the Human Genome Centre, Institute of Medical Science, the University of Tokyo.

## References

1. Albert FW, Kruglyak L. The role of regulatory variation in complex traits and disease. Nat Rev Genet. Nature Publishing Group; 2015;16:197–212.

2. Qi T, Wu Y, Fang H, Zhang F, Liu S, Zeng J, et al. Genetic control of RNA splicing and its distinct role in complex trait variation. Nat Genet. Springer US; 2022;54:1355–63.

3. Dong P, Hoffman GE, Apontes P, Bendl J, Rahman S, Fernando MB, et al. Population-level variation in enhancer expression identifies disease mechanisms in the human brain. Nat Genet. 2022;54:1493–503.

4. Wong JH, Shigemizu D, Yoshii Y, Akiyama S, Tanaka A, Nakagawa H, et al. Identification of intermediate-sized deletions and inference of their impact on gene expression in a human population. Genome Med. Genome Medicine; 2019;11:44.

5. Ashouri S, Wong JH, Nakagawa H, Shimada M, Tokunaga K, Fujimoto A. Characterization of intermediate-sized insertions using whole-genome sequencing data and analysis of their functional impact on gene expression. Hum Genet. Springer Berlin Heidelberg; 2021;140:1201–16. Available from: 10.1007/s00439-021-02291-2

6. Gymrek M, Willems T, Guilmatre A, Zeng H, Markus B, Georgiev S, et al. Abundant contribution of short tandem repeats to gene expression variation in humans. Nat Genet. 2015;48:22–9.

7. Uffelmann E, Huang QQ, Munung NS, de Vries J, Okada Y, Martin AR, et al. Genome-wide association studies. Nat Rev Methods Prim. Springer US; 2021;1. Available from: 10.1038/s43586-021-00056-9

8. Ratnapriya R, Sosina OA, Starostik MR, Kwicklis M, Kapphahn RJ, Fritsche LG, et al. Retinal transcriptome and eQTL analyses identify genes associated with age-related macular degeneration. Nat Genet. 2019;51:606–10.

9. Aragam KG, Jiang T, Goel A, Kanoni S, Wolford BN, Atri DS, et al. Discovery and systematic characterization of risk variants and genes for coronary artery disease in over a million participants. Nat Genet. 2022;54:1803–15.

10. Yao DW, O’Connor LJ, Price AL, Gusev A. Quantifying genetic effects on disease mediated by assayed gene expression levels. Nat Genet. 2020;52:626–33. Available from: 10.1038/s41588-020-0625-2

11. Scotti MM, Swanson MS. RNA mis-splicing in disease. Nat Rev Genet. Nature Publishing Group; 2016;17:19–32.

12. Banday AR, Stanifer ML, Florez-Vargas O, Onabajo OO, Papenberg BW, Zahoor MA, et al. Genetic regulation of OAS1 nonsense-mediated decay underlies association with COVID-19 hospitalization in patients of European and African ancestries. Nat Genet. 2022;54:1103–16.

13. Wu H, Lu Y, Duan Z, Wu J, Lin M, Wu Y, et al. Nanopore long-read RNA sequencing reveals functional alternative splicing variants in human vascular smooth muscle cells. Commun Biol. Springer US; 2023;6:1–12.

14. Kiyose H, Nakagawa H, Ono A, Aikata H, Ueno M, Hayami S, et al. Comprehensive analysis of full-length transcripts reveals novel splicing abnormalities and oncogenic transcripts in liver cancer. PLoS Genet. 2022;18:1–28. Available from: 10.1371/journal.pgen.1010342

15. Sun Q, Han Y, He J, Wang J, Ma X, Ning Q, et al. Long-read sequencing reveals the landscape of aberrant alternative splicing and novel therapeutic target in colorectal cancer. Genome Med. BioMed Central; 2023;15:1–24. Available from: 10.1186/s13073-023-01226-y

16. Veiga DFT, Nesta A, Zhao Y, Mays AD, Huynh R, Rossi R, et al. A comprehensive long-read isoform analysis platform and sequencing resource for breast cancer. Sci Adv. 2022;8:1–14.

17. Yamaguchi K, Ishigaki K, Suzuki A, Tsuchida Y, Tsuchiya H, Sumitomo S, et al. Splicing QTL analysis focusing on coding sequences reveals mechanisms for disease susceptibility loci. Nat Commun. Springer US; 2022;13:1–13.

18. Glinos DA, Garborcauskas G, Hoffman P, Ehsan N, Jiang L, Gokden A, et al. Transcriptome variation in human tissues revealed by long-read sequencing. Nature. Springer US; 2022.

19. Auton A, Abecasis GR, Altshuler DM, Durbin RM, Bentley DR, Chakravarti A, et al. A global reference for human genetic variation. Nature. 2015;526:68–74.

20. Li H. Minimap2: Pairwise alignment for nucleotide sequences. Bioinformatics. 2018;34:3094–100.

21. Purcell S, Neale B, Todd-Brown K, Thomas L, Ferreira MAR, Bender D, et al. PLINK: A tool set for whole-genome association and population-based linkage analyses. Am J Hum Genet. 2007;81:559–75.

22. Kadri NK, Mapel XM, Pausch H. The intronic branch point sequence is under strong evolutionary constraint in the bovine and human genome. Commun Biol. Springer US; 2021;4:1–13.

23. Birney E, Clamp M, Durbin R. GeneWise and Genomewise. Genome Res. 2004;14:988–95. Available from: www.genome.org

24. Khan A, Zhang X. dbSUPER: a database of super-enhancers in mouse and human genome. Nucleic Acids Res. 2016;44. Available from: http://bioinfo.au.

25. ENCODE Project Consortium. An integrated encyclopedia of DNA elements in the human genome ENCODE Encyclopedia of DNA Elements. Nature. 2012;488. Available from: http://encodeproject.org/ENCODE/

26. Shabalin AA. Matrix eQTL: Ultra fast eQTL analysis via large matrix operations. Bioinformatics. 2012;28:1353–8. Available from: http://www.bios.unc.edu/research/genomic_software/Matrix_eQTL

27. Benner C, Spencer CCA, Havulinna AS, Salomaa V, Ripatti S, Pirinen M. FINEMAP: Efficient variable selection using summary data from genome-wide association studies. Bioinformatics. 2016;32:1493–501. Available from: http://www.christianbenner.com.

28. Strober BJ, Wen X, Wucher V, Kwong A, Lappalainen T, Li X, et al. The GTEx Consortium atlas of genetic regulatory effects across human tissues. Science (80- ). 2020;18:1318–30.

29. Buniello A, Macarthur JAL, Cerezo M, Harris LW, Hayhurst J, Malangone C, et al. The NHGRI-EBI GWAS Catalog of published genome-wide association studies, targeted arrays and summary statistics 2019. Nucleic Acids Res. Oxford University Press; 2019;47:D1005–12.

30. Jaganathan K, Kyriazopoulou Panagiotopoulou S, McRae JF, Darbandi SF, Knowles D, Li YI, et al. Predicting Splicing from Primary Sequence with Deep Learning. Cell. Elsevier; 2019;176:535–548.e24. Available from: 10.1016/j.cell.2018.12.015

31. Eddy SR. Accelerated profile HMM searches. PLoS Comput Biol. 2011;7:1002195. Available from: www.ploscompbiol.org

32. Chandra V, Bhattacharyya S, Schmiedel BJ, Madrigal A, Gonzalez-Colin C, Fotsing S, et al. Promoter-interacting expression quantitative trait loci are enriched for functional genetic variants. Nat Genet. 2021;53:110–9. Available from: 10.1038/s41588-020-00745-3

33. Agirre E, Oldfield AJ, Bellora N, Segelle A, Luco RF. Splicing-associated chromatin signatures: a combinatorial and position-dependent role for histone marks in splicing definition. Nat Commun. 2021;12:1–16.

34. Ward LD, Kellis M. HaploReg v4: Systematic mining of putative causal variants, cell types, regulators and target genes for human complex traits and disease. Nucleic Acids Res. 2016;44:D877–81. Available from: https://academic.oup.com/nar/article/44/D1/D877/2503117

35. Phanstiel DH, Van Bortle K, Spacek D, Hess GT, Shamim MS, Machol I, et al. Static and Dynamic DNA Loops form AP-1-Bound Activation Hubs during Macrophage Development. Mol Cell. Elsevier Inc.; 2017;67:1037–1048.e6. Available from: 10.1016/j.molcel.2017.08.006

36. Mucaki EJ, Shirley BC, Rogan PK. Expression Changes Confirm Genomic Variants Predicted to Result in Allele-Specific, Alternative mRNA Splicing. Front Genet. 2020;11:1–16.

37. Connally N, Nazeen S, Lee D, Shi H, Stamatoyannopoulos J, Chun S, et al. The missing link between genetic association and regulatory function. Elife. 2022;11:1–35.

38. Haralambieva IH, Ovsyannikova IG, Kennedy RB, Larrabee BR, Zimmermann MT, Grill DE, et al. Genome-wide associations of CD46 and IFI44L genetic variants with neutralizing antibody response to measles vaccine. Hum Genet. Springer Berlin Heidelberg; 2017;136:421–35.

39. Akiyama M, Ishigaki K, Sakaue S, Momozawa Y, Horikoshi M, Hirata M, et al. Characterizing rare and low-frequency height-associated variants in the Japanese population. Nat Commun. 2019;10. Available from: 10.1038/s41467-019-12276-5

40. Karlsson Linnér R, Biroli P, Kong E, Meddens SFW, Wedow R, Fontana MA, et al. Genome-wide association analyses of risk tolerance and risky behaviors in over 1 million individuals identify hundreds of loci and shared genetic influences. Nat Genet. 2019;51:245–57.

41. Alarcón-Riquelme ME, Ziegler JT, Molineros J, Howard TD, Moreno-Estrada A, Sánchez-Rodríguez E, et al. Genome-Wide Association Study in an Amerindian Ancestry Population Reveals Novel Systemic Lupus Erythematosus Risk Loci and the Role of European Admixture. Arthritis Rheumatol. 2016;68:932–43.

42. Clark AR, Ohlmeyer M. Protein phosphatase 2A as a therapeutic target in inflammation and neurodegeneration. Pharmacol Ther. Elsevier Inc.; 2019;201:181–201. Available from: 10.1016/j.pharmthera.2019.05.016

