## Additional file1 for "Long-read sequencing reveals novel isoform-specific eQTLs and regulatory mechanisms of isoform expression"

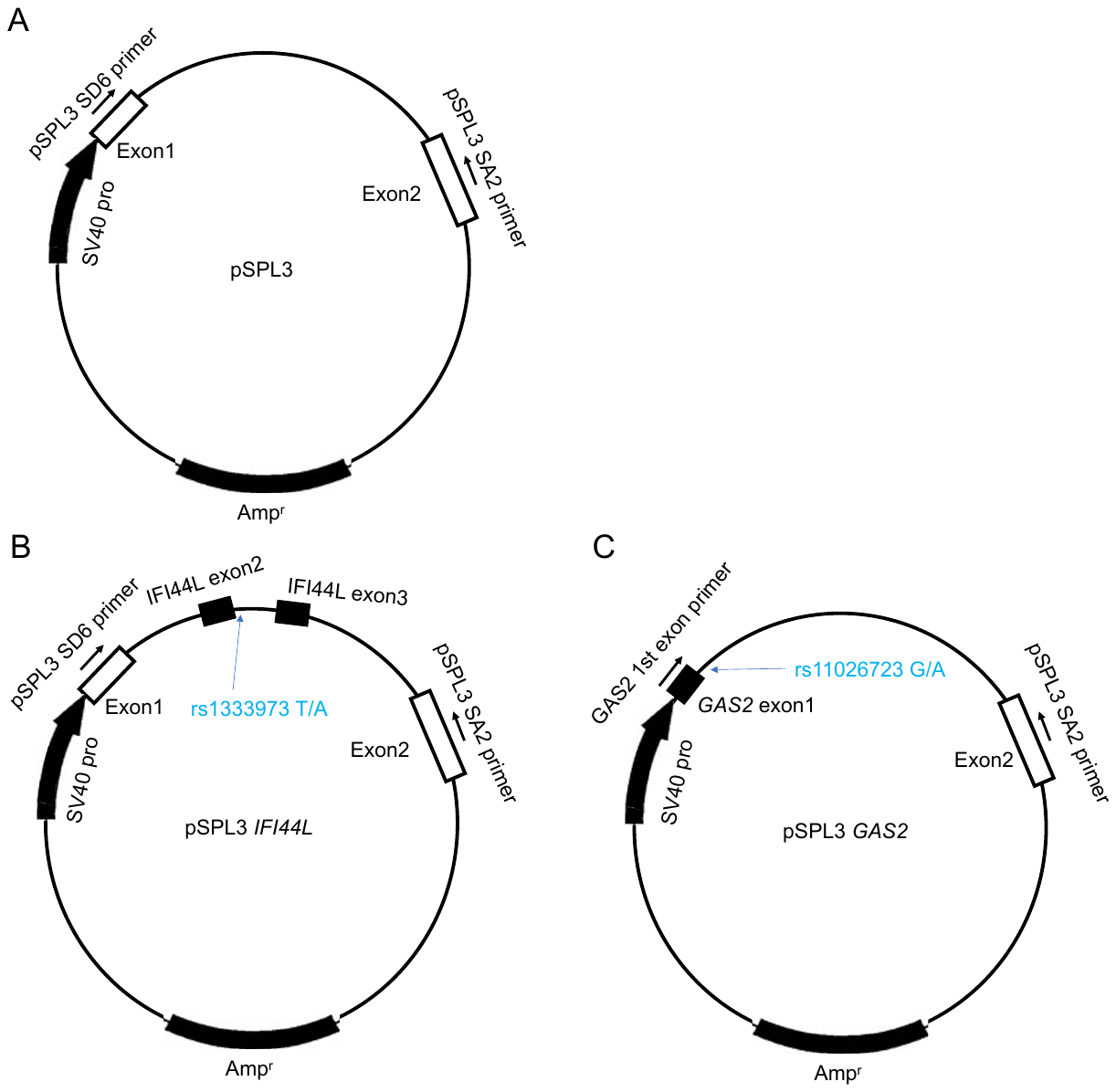

**Figure S1** Structure of vectors for minigene splicing assays.

(A) Structure of pSPL (empty) vector.
(B) Structure of pSPL *IFI44L* vector.
(C) Structure of pSPL *GAS2* vector.

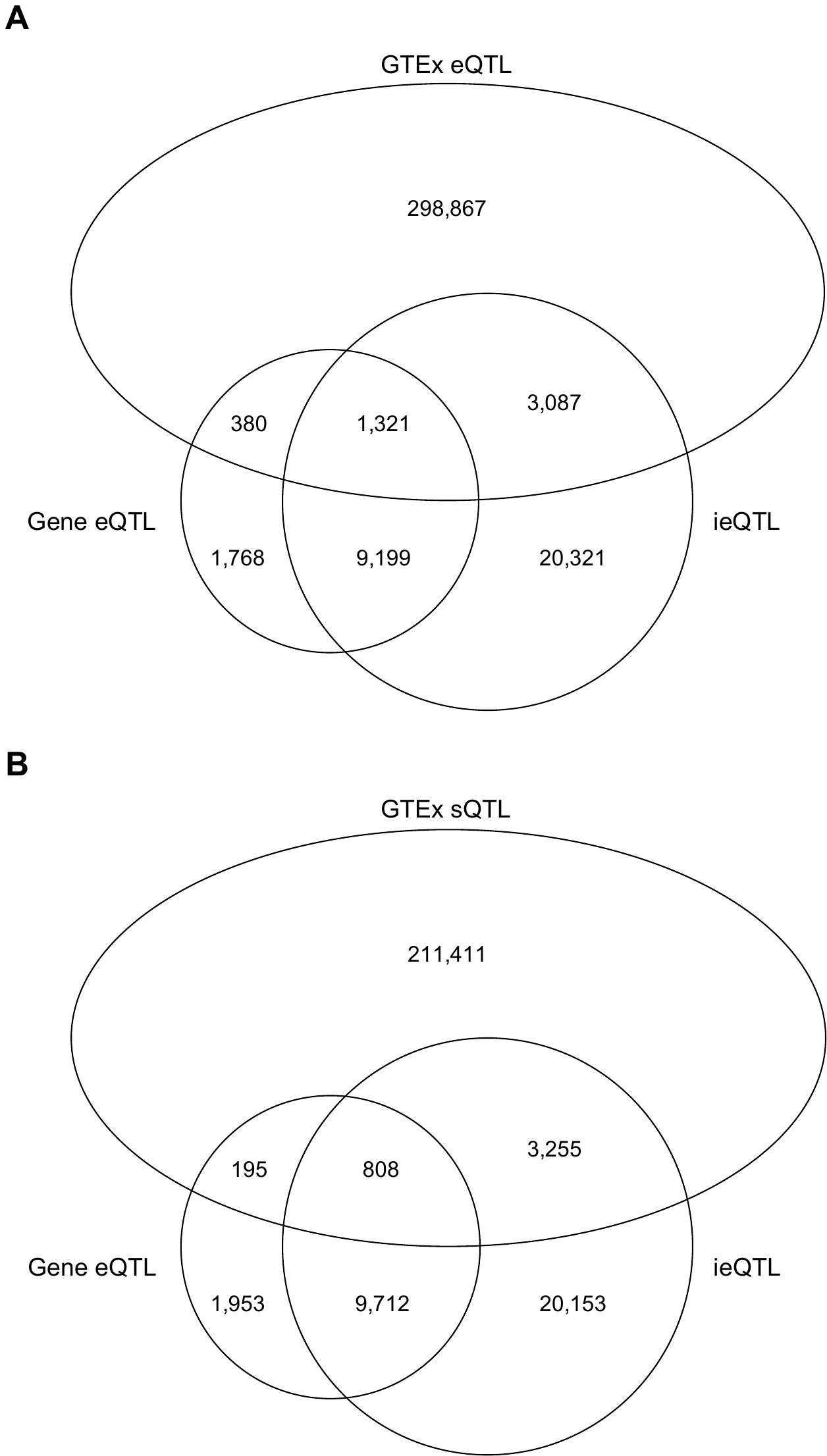

**Figure S2** Comparison with eQTL identified by GTEx project.

(A) GTEx eQTL.

(B) GTEx sQTL.

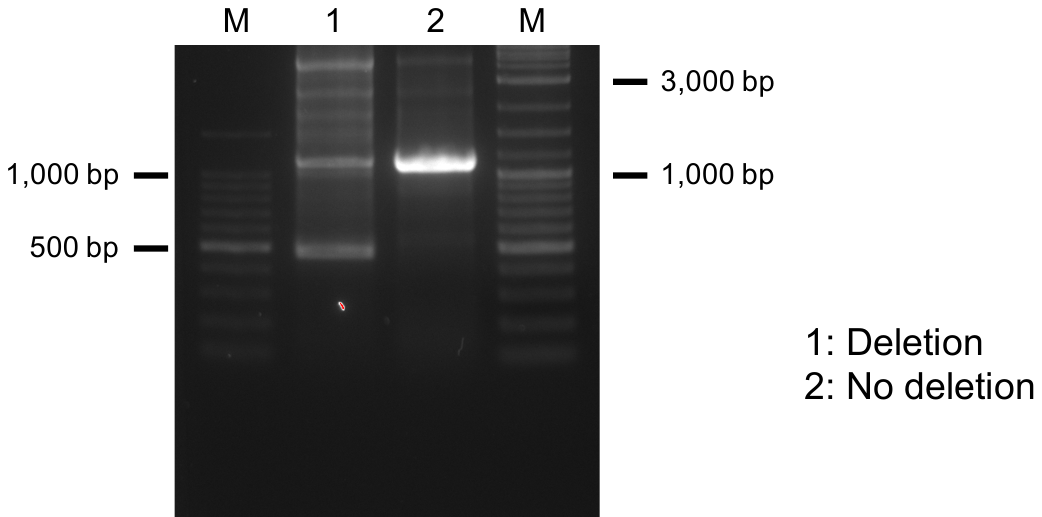

**Figure S3** Deletion generated in HEK293T cells using CRISPR-Cas9 system.

M: marker. Lane1: HEK293T cells with chr10:103,342,876-103,343,562 deletion. Lane2: HEK293T cells without the deletion.

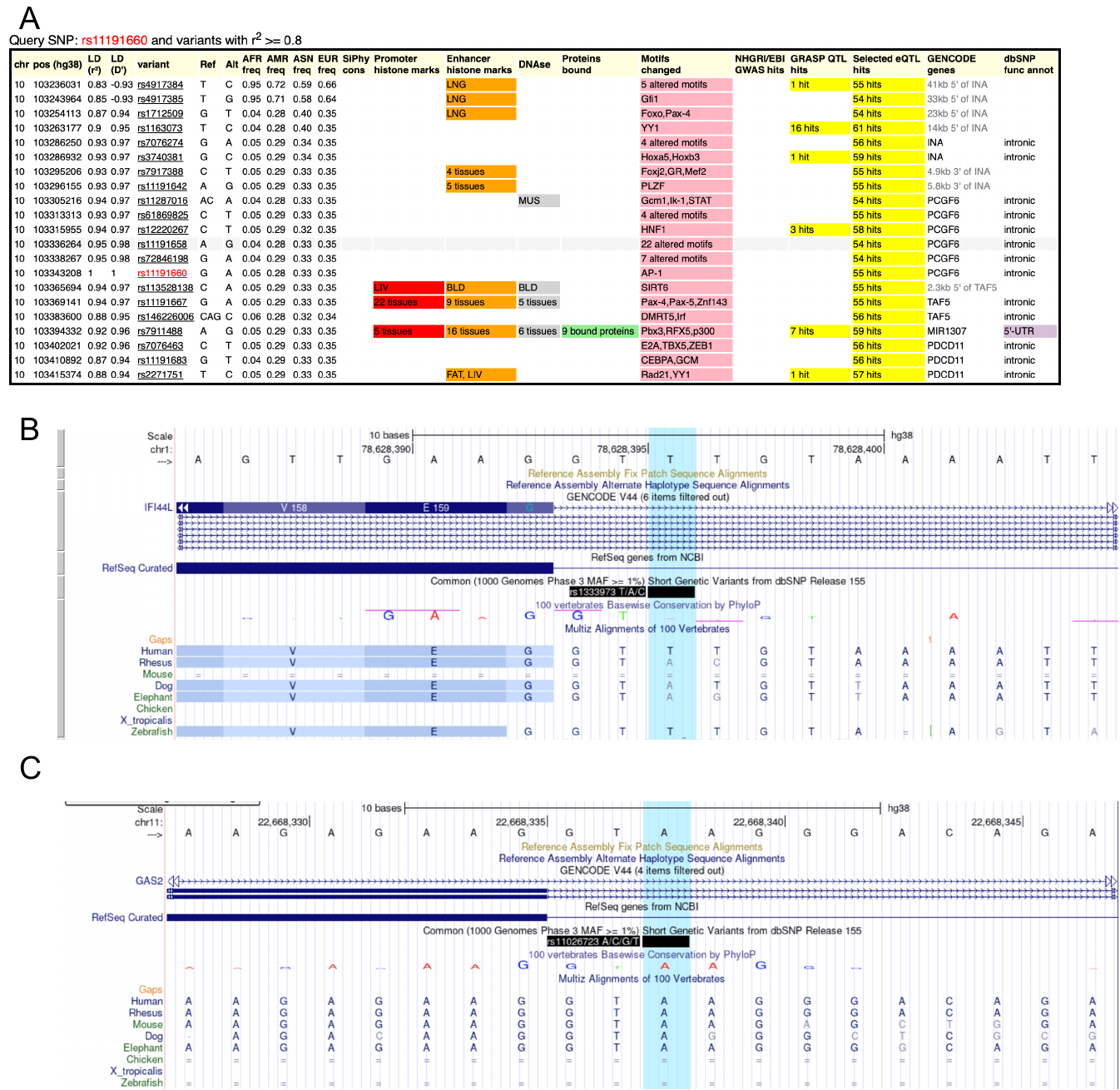

**Figure S4** Feature of SNPs.

(A) Analysis of rs11191660 using HaploReg website.
(B) Evolutionary conservation of rs1333973. The position of rs1333973 was not highly conserved.
(C) Evolutionary conservation of rs11026723. The position of rs11026723 was highly conserved.
